# Sex and education differences in trajectories of physiological ageing: longitudinal analysis of a prospective English cohort study

**DOI:** 10.1101/2025.01.06.25320036

**Authors:** Mikaela Bloomberg, Andrew Steptoe

## Abstract

**Background:** Physiological age (PA) derived from clinical indicators including blood-based biomarkers and tests of physiological function can be compared with chronological age to examine disparities in health between older adults of the same age. Though education interacts with sex to lead to inequalities in healthy ageing, their combined influence on longitudinally-measured PA has not been explored. We derived PA based on longitudinally-measured clinical indicators and examined how sex and education interact to inform PA trajectories.

**Methods:** Three waves of clinical indicators (2004/05-2012/13) drawn from the English Longitudinal Study of Ageing (ages 50-100 years) were used to estimate PA, which was internally validated by confirming associations with incident chronic conditions, functional limitations, and memory impairment after adjustment for chronological age and sex. Joint models were used to construct PA trajectories in 8,891 ELSA participants to examine sex and educational disparities in PA.

**Findings:** Among the least educated participants, there were negligible sex differences in PA until age 60 (sex difference [men-women] age 50=-0.6 years [95% confidence interval=-2.2-0.6]; age 60=0.4 [-0.6-1.4]); at age 70, women were 1.5 years (0.7-2.2) older than men. Among the most educated participants, women were 3.8 years (1.6-6.0) younger than men at age 50, and 2.7 years (0.4-5.0) younger at age 60, with a non-significant sex difference at age 70.

**Interpretation:** Higher education provides a larger midlife buffer to physiological ageing for women than men. Policies to promote gender equity in higher education may contribute to improving women’s health across a range of ageing-related outcomes.

## Introduction

It is well-documented that the central ageing processes driving pathophysiological changes observed during ageing are not perfectly correlated with chronological age (1, 2), leading to considerable heterogeneity in health between older adults of the same age. Biological ageing metrics based on telomere length or DNA methylation attempt to explain variability in ageing-related disease better than chronological age alone (3–7). However, biological age derived from clinical indicators including blood-based biomarkers and tests of physiological function (6–11)—frequently referred to as physiological or phenotypic age—may explain variation in health outcomes better than molecular or cellular measures, because these indicators reflect physiological changes more closely related to health outcomes (8, 10). Where biological age is intended to be a universal measure of cellular ageing, physiological age captures downstream effects of the ageing process on multiple organ systems to serve as a healthy ageing index. Physiological age can then be compared with chronological age to examine characteristics of individuals with physiological age younger or older than their chronological age and can thus be used to identify socioeconomic disparities in healthy ageing.

Among commonly examined socioeconomic drivers of inequalities in healthy ageing, education is of particular importance due to its propagating effects on socioeconomic position during the life course, influencing a myriad of health-related factors including access to healthcare, health behaviours, and health literacy, and culminating in its impact on late life morbidity and mortality (12, 13). Education also has key interactions with gender—which is commonly informed by sex—as historically, women were restricted in access to education and limited to lower levels of educational attainment (14). While there is a substantial body of literature examining how interactions of sex and education impact individual health outcomes, and how sex and education are independently associated with metrics of biological age, how sex and education combine to influence physiological ageing overall is not yet clear. Previous studies examining either sex or education disparities in biological age are largely cross-sectional (15–23). Studies examining sex differences in physiological age produced using longitudinally-measured clinical indicators are undertaken in small, highly-selected study populations (N=179-802) (24–26).

To examine how sex and education interact to inform physiological ageing, we derived a physiological ageing measure based on clinical indicators assessed at three time points (2004/05-2012/13). We examined sex and educational differences in longitudinal PA trajectories from 8,891 participants aged 50-100 years from the English Longitudinal Study of Ageing (ELSA).

## Methods

### Data sources

ELSA is a nationally-representative cohort study of the English population aged ≥50 years with biennial data collection beginning in 2002/03 continuing until 2018/19; the most recent wave of data collection occurred in 2021/23. Details of survey design are available elsewhere (27). During waves 2 (2004/05), 4 (2008/09), and 6 (2012/13), ELSA participants underwent a series of physiological assessments and had blood samples drawn during a home visit from a study nurse. ELSA participants with at least one nurse visit were eligible for inclusion in analyses. Waves 2-10 of ELSA (2004/05-2021/23) were also used to internally validate physiological age (Appendix 2a).

### Sex, education, and covariates

Sex and education were self-reported at the first wave of biomarker collection. Participants were asked to report their “sex” as “man” or “woman”. ELSA survey materials do not delineate between sex and gender; sex was referred to in the survey and is therefore used in the present study. Participants were also asked about their highest educational qualification, categorised into less than high school (low education), high school (intermediate education), or above high school (high education).

Participants were also asked to report their birth year which was categorised into seven-year birth cohorts (1911-1962), chosen to maximise the age range in each birth cohort whilst minimising birth cohort effects.

### Derivation and internal validation of physiological age

Clinical indicators collected during nurse visits in waves 2, 4, and 6 included those pertaining to the cardiovascular system (pulse pressure, systolic blood pressure, diastolic blood pressure, mean arterial pressure), respiratory system (forced vital capacity [FVC], forced expiratory volume in one second [FEV]), the haematologic system (haemoglobin concentration, fibrinogen, C-reactive protein [CRP], ferritin), metabolism (fasting glucose, glycated haemoglobin, total cholesterol, LDL cholesterol, HDL cholesterol, triglyceride), and muscle and fat (grip strength, waist circumference). Procedures for indicator collection and processing are reported in Appendix 1a.

Derivation of physiological age was performed using the principal component analysis (PCA) method, a commonly used method of physiological age estimation (28–31); details are available in Appendix 1b. In line with previous studies, physiological age was derived using 822 ELSA participants who did not report chronic conditions (Appendix 3a) and was then calculated for all participants in the main analytic sample. Clinical indicators selected using the PCA method for inclusion in physiological age were pulse pressure, systolic blood pressure, fibrinogen, CRP, glycated haemoglobin, FEV, FVC, and grip strength (Appendix 3b). These indicators were used to produce a physiological score which was converted to a physiological age in years using previously described methods (28–31) (Appendix 1b). Finally, we estimated ageing acceleration (physiological age – chronological age).

We first described characteristics of men and women in the analytic sample using *t*-test and Pearson’s *x*^2^ test for continuous and categorical variables respectively. To internally validate physiological age, we used Cox proportional hazards models adjusted for chronological age and sex to examine associations between each individual’s first measurement of ageing acceleration (occurring at either wave 2, 4, or 6) and incidence of the following ageing-related outcomes occurring during waves 2-10 of ELSA: 1) functional limitations in activities of daily living (ADL), instrumental activities of daily living (IADL), and mobility activities; 2) memory impairment; and 3) ageing-related chronic conditions. Details are available in Appendix 1c.

### Trajectories of physiological age

We used joint models to examine trajectories of physiological age. These models simultaneously estimate a longitudinal sub-model (linear mixed effects model with a chronological age time scale and a random intercept and slope at the individual level) and a survival sub-model (Weibull model) which are linked through shared random effects and are used to account for data missing-not-at-random in the longitudinal process (32).

Model 1 included chronological age centred at age 50 (*CA*)*, birth cohort*, and the interaction between birth cohort and CA (denoted *birth cohort x CA*). Visual inspection using local polynomial regression suggested the relationship between physiological age and CA was linear (Appendix 2b); we therefore did not include higher-order CA terms. Birth cohort was included as a continuous variable centred at the 1939-1945 cohort. To examine sex differences in physiological ageing, we added *sex* and *sex x CA* to Model 1 to yield Model 2. For Model 3, we added *education* and *education x CA* to Model 2 to examine educational differences in physiological ageing. Finally, for Model 4, we added *sex x education* and *sex x education x CA* to Model 3 to determine whether sex differences in physiological age differed by education level.

We used these models to plot trajectories of physiological age from chronological ages 50-80 years in men and women (Model 2), by education level (Model 3), and in men and women by education level (Model 4). This age range was chosen to reduce birth cohort effects. We reported sex and education differences at ages 50, 60, and 70 and again after eight years of follow-up (the maximum follow-up period; corresponding to the difference at ages 58, 68, and 78). Analyses were performed in StataMP 18.0 or R 4.2.2 with a two-sided p<0.050 considered significant.

## Results

Of 12,291 ELSA respondents aged ≥ 50 years with at least one nurse visit during waves 2, 4, or 6, 8,906 (72.4%) had complete biomarker data. Of these 8,906, 15 (0.2%) were missing education and were excluded, leading to 8,891 participants retained for the analyses (Appendix 2c). Table 1 shows characteristics of the analytic sample. Of 8,891 participants, 4094 (46.1%) were men and 4797 (53.1%) were women. Men and women had the same mean chronological age (64.1 years, SD=9.1 for men or 9.4 for women; p=0.94). Men had younger physiological ages than women (mean=68.4 years [SD=18.4] for men, mean=69.2 [SD=20.4] for women; p=0.045). Men had physiological ages on average 4.2 years (SD=13.1) older than their chronological age compared with 5.1 years (SD=14.3) for women (p=0.0048). Men were more educated than women (p<0.0001).

**Table 1.** Characteristics of the analytic sample at first physiological age measurement.

|  | **Men**<br>N=4094 | **Women**<br>N=4797 | P-value |
| --- | --- | --- | --- |
| Chronological age, mean (SD) | 64.1 (9.1) | 64.1 (9.4) | 0.94 |
| Physiological age, mean (SD) | 68.4 (18.7) | 69.2 (20.4) | 0.045 |
| Ageing acceleration, mean (SD) | 4.2 (13.1) | 5.1 (14.3) | 0.0048 |
| Highest educational qualification |  |  |  |
| Less than high school | 1586 (38.7) | 2198 (45.8) |  |
| High school | 1849 (45.2) | 2107 (43.9) | <0.0001 |
| Above high school | 659 (16.1) | 492 (10.3) |  |

N (%) shown unless otherwise indicated. Abbreviations: SD, standard deviation.

**Table 2.** Associations between ageing acceleration and ageing-related outcomes.

|  | **HR for 5-year change in**<br>**ageing acceleration (95% CI)** | *P-value* |
| --- | --- | --- |
| *Functional limitations* |  |  |
| ADL | 1.15 (1.14-1.17) | <0.0001 |
| IADL | 1.16 (1.15-1.17) | <0.0001 |
| Mobility activities | 1.09 (1.09-1.10) | <0.0001 |
| Memory impairment | 1.16 (1.14-1.17) | <0.0001 |
| *Chronic conditions* |  |  |
| Diabetes | 1.34 (1.30-1.38) | <0.0001 |
| Lung disease | 1.23 (1.19-1.28) | <0.0001 |
| Cardiovascular disease | 1.09 (1.06-1.11) | <0.0001 |
| Stroke | 1.16 (1.12-1.21) | <0.0001 |
| High cholesterol | 1.08 (1.06-1.09) | <0.0001 |
| High blood pressure | 1.26 (1.23-1.28) | <0.0001 |
| Cancer | 1.03 (1.00-1.05) | 0.065 |
| Arthritis | 1.06 (1.03-1.08) | <0.0001 |
| Osteoporosis | 1.04 (1.01-1.07) | 0.018 |
| Dementia | 1.09 (1.05-1.14) | <0.0001 |
| Parkinson's disease | 0.98 (0.90-1.07) | 0.63 |

Based on Cox proportional hazards models adjusted for sex and chronological age.
Outcomes ascertained from waves 2-10 of ELSA (2004/05-2021/23).
Abbreviations: HR, hazard ratio; CI, confidence interval; ADL, activities of daily living; IADL, instrumental activities of daily living.

### Internal validation of physiological age

Table 1 shows hazard ratios (HR) for associations between ageing acceleration and incident ageing-related outcomes occurring between 2004/05-2021/23. The mean follow-up period for these analyses was 9.5 years (SD=5.5) for men or 9.9 years (SD=5.4) for women. After adjusting for sex and chronological age, a five-year increase in ageing acceleration (physiological age – chronological age) was associated with increased incidence of ADL (HR=1.15 [95% CI=1.14-1.17]), IADL (1.16 [1.15-1.17]), and mobility limitations (1.09 [1.09-1.10]), memory impairment (1.16 [1.14-1.17]), diabetes (1.34 [1.30-1.38]), lung disease (1.23 [1.19-1.28]), cardiovascular disease (1.09 [1.06-1.11]), stroke (1.16 [1.12-1.21]), high cholesterol (1.08 [1.06-1.09]), high blood pressure (1.26 [1.23-1.28]), arthritis (1.06 [1.03-1.08]), dementia (1.09 [1.05-1.14]; p<0.0001 for all), and osteoporosis (1.04 [1.01-1.07]; p=0.018). There was no association between ageing acceleration and cancer (1.03 [1.00-1.05]; p=0.065) or Parkinson’s disease (0.98 [0.90-1.07]; p=0.63).

### Trajectories of physiological age

The mean follow-up duration for physiological age trajectories was the same for men and women (mean=2.6 years [SD=3.1]). After adjusting for birth cohort, participants had an average physiological age of 49.4 years (48.7-50.2) at age 50 years, 63.2 (62.8-63.6) at age 60, and 77.0 (76.6-77.4) at age 70. Over eight years of follow-up, physiological age increased on average 11.0 years (10.7-11.4).

Physiological ages of women increased faster than men (p<0.0001); during eight years of follow-up, women aged 1.0 year (0.6-1.4) more than men (Figure 1). Women were physiologically 0.9 years (0.0-1.8) younger than men at age 50 but had similar ages to men at age 60 (0.3 years [-0.2-0.9] older than men) and were 1.6 years (1.0-2.1) older than men at age 70. At the end of eight years of follow-up, women were 0.9 years [-0.5-0.7] older than men when aged 50 years at baseline (though this difference did not reach statistical significance), 1.3 (0.8-1.8) older when aged 60 at baseline, or 2.5 (1.8-3.3) older when aged 70 at baseline.

**Figure 1.**
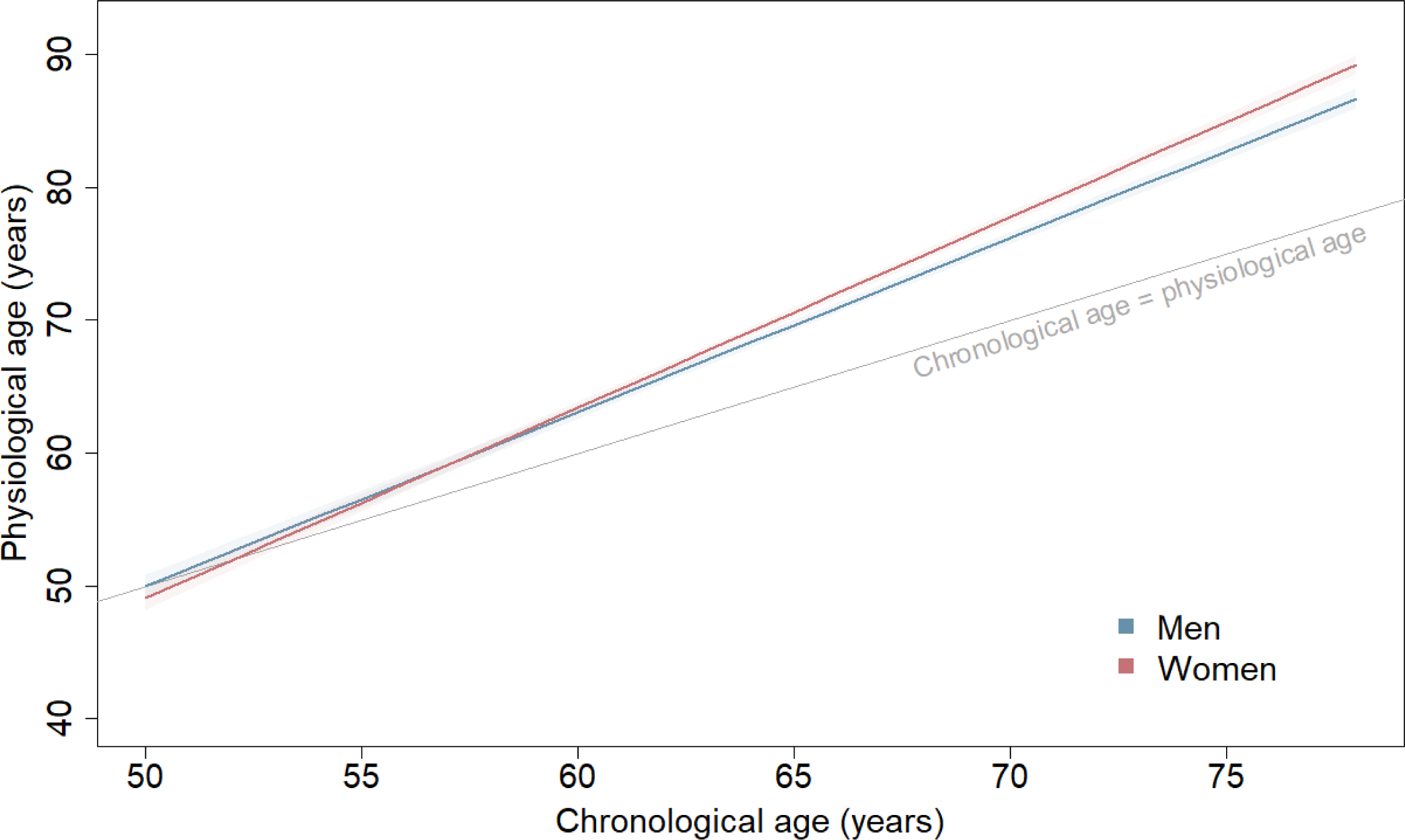
Physiological ageing trajectories from age 50 to 80 years in men and women. Average physiological age plotted against chronological ages 50 to 80 years in men and women. Based on joint model including chronological age, birth cohort, sex, and interactions of 1) birth cohort and chronological age; and 2) sex and chronological age. Plotted for reference values of covariates (1939-1945 birth cohort).

At first physiological age measurement, participants in the low education group had physiological ages on average 9.3 years (SD=13.7) older than their chronological age, compared with 2.1 years (SD=12.8) older in the intermediate education group. Participants in the high education group had physiological ages on average 1.7 years (SD=12.3) younger than their chronological age.

After adjusting for birth cohort and sex, participants in the intermediate education group had physiological ages 4.7 years (3.7-5.7) younger than the low education group at age 50, 4.2 (3.6-4.9) younger at age 60, or 3.7 (3.2-4.3) younger at age 70 (Figure 2). Participants in the high education group had physiological ages 7.6 years (6.2-9.0) younger than the low education group at age 50, 7.3 (6.4-8.1) younger at age 60, and 6.9 (6.0-7.9) younger at age 70.

**Figure 2.**
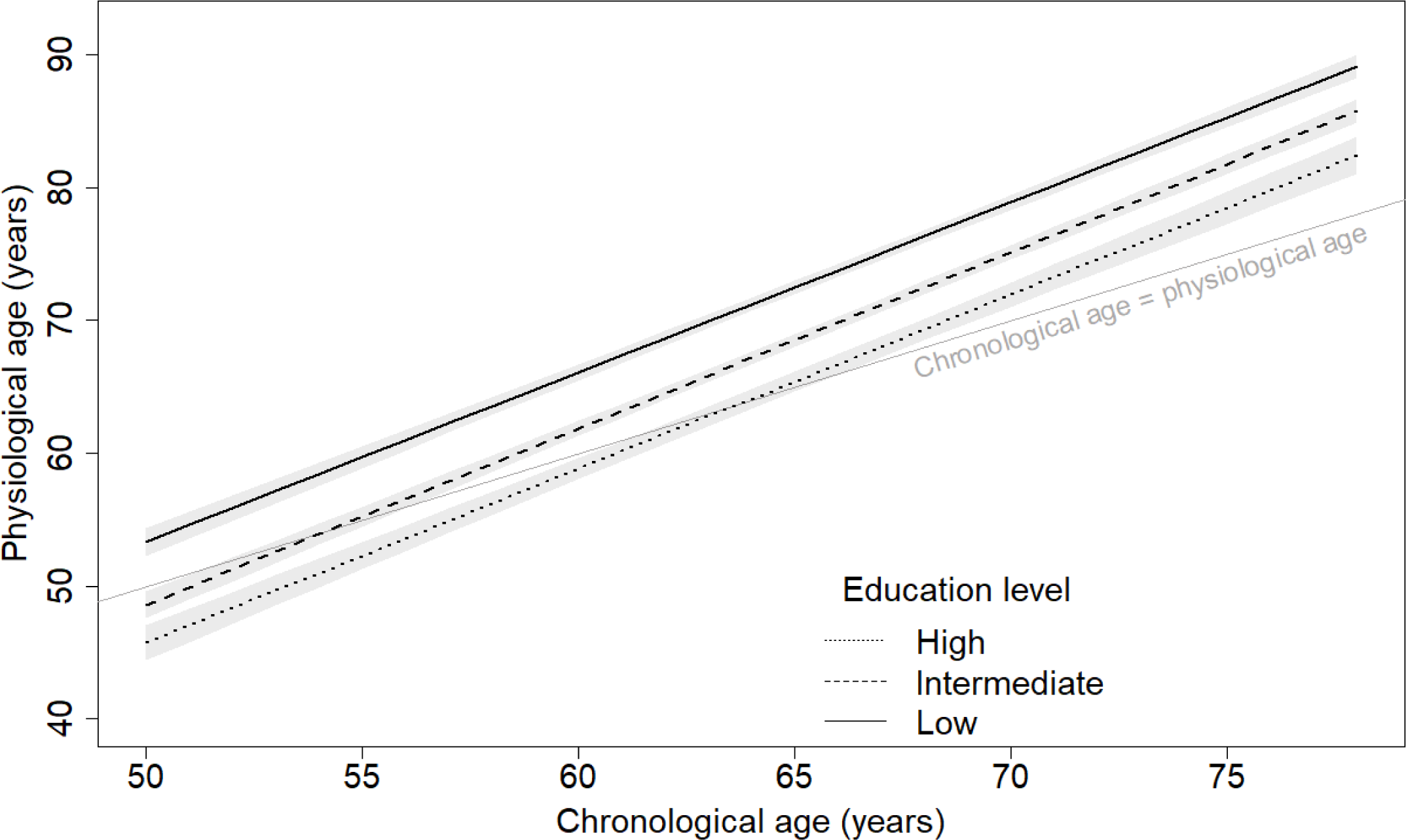
Physiological ageing trajectories from age 50 to 80 years by education level. Physiological age plotted against chronological ages 50 to 80 by education level (low=below high school; intermediate=high school; high=above high school). Based on joint model including chronological age, birth cohort, sex, education and interactions of 1) birth cohort and chronological age; 2) sex and chronological age; and 3) education and chronological age. Plotted for reference values of covariates (men, 1939-1945 birth cohort).

There were negligible differences in the pace of physiological ageing between education categories (p=0.20). Over eight years of follow-up, participants with low education had physiological ages that increased 10.2 years (9.7-10.7), compared with 10.6 (10.2-11.1) for intermediate education and 10.5 (9.8-11.1) for high education.

In general, sex differences in physiological ageing were similar for low and intermediate education levels (p*_intermediate education x sex_*=0.68; Figure 3) and sex differences in the pace of physiological ageing did not vary across levels of education (p*_intermediate education x sex x CA_*=0.97; p*_high education x sex x CA_*=0.37). However, sex differences in physiological age in the high education group differed from the low education group (p*_high education x sex_*=0.021). In the low education group, women had similar physiological ages to men at age 50 (0.6 years [-0.9-2.2] younger than men) and 60 (0.4 years [-0.6-1.4] older than men), and were 1.5 years (0.7-2.2) older than men at age 70 years; sex differences were similar in the intermediate education group. By contrast, in the high education group, women had physiological ages 3.8 years (1.6-6.0) younger than men at age 50, 2.7 years (0.4-5.0) younger than men at age 60, and 1.7 (−0.9-4.3) years younger than men at age 70, though the sex difference at age 70 did not reach statistical significance.

**Figure 3.**
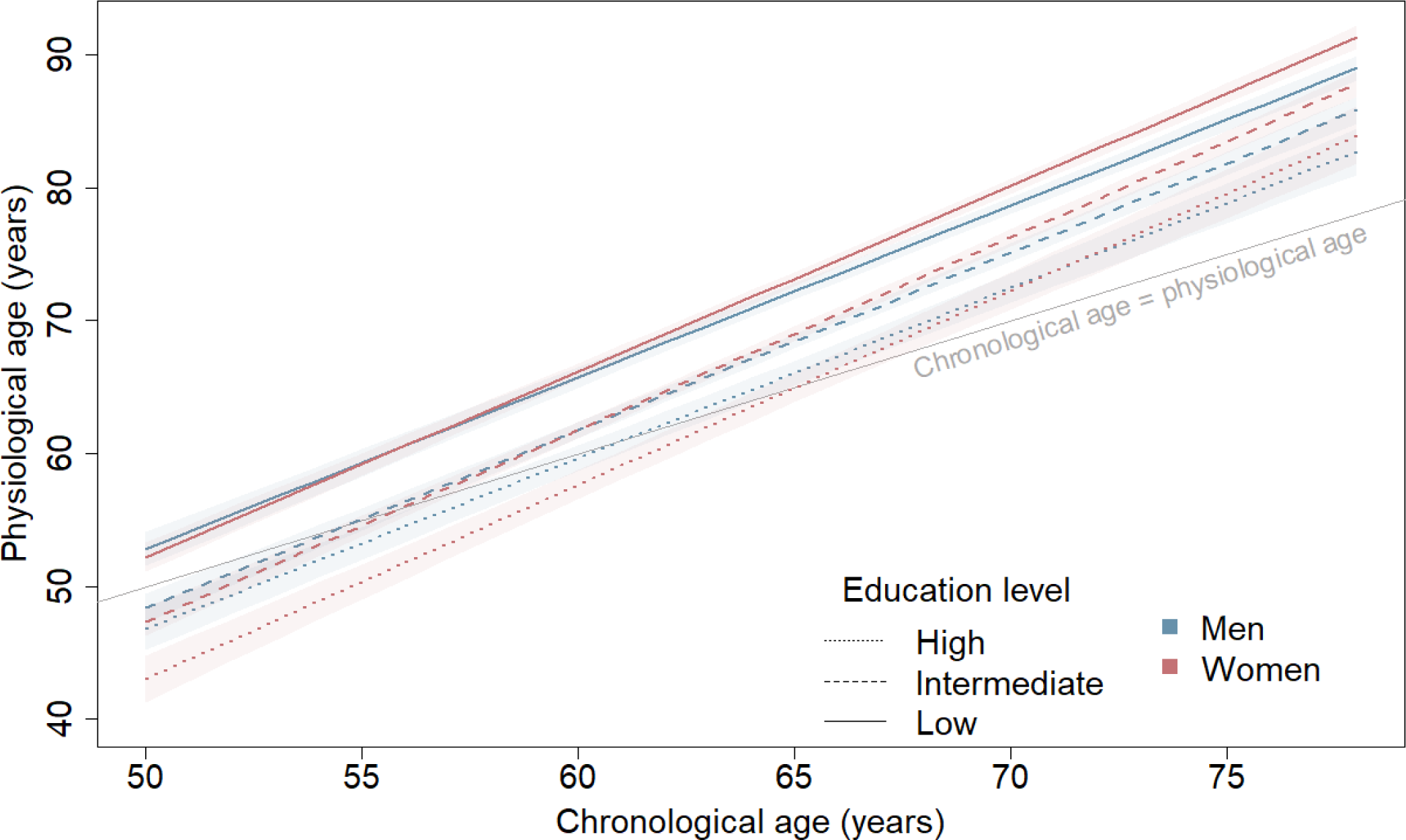
Physiological ageing trajectories from age 50 to 80 years in men and women by education level. Physiological age plotted against chronological ages 50 to 80 by sex and education level (low=below high school; intermediate=high school; high=above high school). Based on joint model including chronological age, birth cohort, sex, education and interactions of 1) birth cohort and chronological age; 2) sex and chronological age; 3) education and chronological age; 4) sex and education; and 5) sex, education, and chronological age. Plotted for reference values of covariates (1939-1945 birth cohort).

## Discussion

After deriving a physiological ageing measure and examining physiological ageing trajectories in 8,891 adults aged 50-100 years, we found key sex and educational differences in physiological ageing. First, before taking education into account, sex differences in physiological age were minor before chronological age 60 but grew progressively larger as women aged faster than men. Second, more education was associated with lower physiological age but no difference in the pace of physiological ageing. Finally, examination of interactions between sex and education revealed that high education provided a midlife benefit for women, such that women educated above high school level were physiologically younger than men until chronological age 70. By contrast, women educated to high school level or below had physiological ages similar to men between chronological ages 50 and 60 and physiological ages increasingly older than men from age 60 onward. These results suggest education above high school level may be important to reduce female disadvantages in physiological ageing.

The main strengths of this study include its longitudinal measure of physiological age, broad age and birth year range, with data drawn from a large and nationally-representative study population. Previous cross-sectional studies examining sex and educational disparities in physiological ageing may be influenced by birth cohort effects, wherein systemic improvements in factors such as socioeconomic conditions and healthcare lead to overestimating physiological age at older chronological ages. The broad age range—where ages are represented in multiple birth cohorts— allowed us to consider birth cohort effects in our analysis. Our physiological ageing measure was derived using routinely-collected clinical indicators, and can therefore easily be derived in other studies or clinical settings. Our physiological ageing measure was also associated with a range of ageing-related health outcomes pertaining to multiple organ systems, indicating it is an internally valid holistic measure of ageing. Finally, we accounted for differential attrition in the analytic sample by simultaneously modelling attrition using joint models.

This study has several limitations. We were limited in choice of clinical indicators to those routinely collected during nurse visits in ELSA; for example, while biological clocks like DunedinPACE distil the longitudinally-measured clinical biomarker-based Pace of Aging down to a single DNA-methylation blood test (10), the data needed to produce DunedinPACE are not available in ELSA. We were unable to externally validate our measure of physiological ageing because comparable longitudinally-measured clinical indicators are not yet widely available in other similar national cohort studies of ageing, so we emphasise that the physiological ageing measure produced in the present study is intended as an internally-validated healthy ageing index. Areas of future research include development of a longitudinal measure of physiological age that can be applied in multiple diverse study populations to compare health disparities across populations.

While previous findings using epigenetic clocks suggest minor sex differences or lower biological ages in women (18–23), our results are consistent with studies using physiological age derived based on clinical indicators which generally suggest that women have higher physiological ages than men (24–26). This evidence supports the adage that women ‘live longer in worse health’ than men, where lower epigenetic age corresponds to greater longevity and older physiological ages are consistent with experiencing worse health. We also found that women aged faster than men. Previous studies examining sex differences in longitudinal trajectories of physiological ageing give mixed results (24–26). One study was undertaken in 179 healthy Japanese men and women aged 30-77 years at baseline and found women aged slightly slower than men but had higher physiological ages at baseline (24); two studies were undertaken in up to 802 participants aged 45-85 at baseline from the Swedish Adoption/Twin Study of Aging to find negligible sex differences in the pace of physiological ageing but higher physiological ages in women (25, 26). Our study expands on this previous evidence undertaken in highly selected study populations, as it examines physiological ageing using data draw from a large, nationally-representative study population that includes participants reporting chronic conditions.

Our findings are in line with previous studies that suggest more education is associated with younger biological age (15–17), and studies that suggest gender disparities in cognitive and functional ageing may occur due to gender differences in socioeconomic characteristics (34, 35). Health benefits of education include access to healthcare, improved health literacy, and engagement in healthy behaviours (36). We also examined interactions between sex and education to show that education above high school level afforded a larger benefit to physiological age for women compared with men. This is consistent with a large body of literature which finds education may confer greater health benefits for women than for men, as women otherwise have fewer socioeconomic resources such as power, authority, or earnings (37). Furthermore, while more education is associated with fewer unhealthy behaviours such as excessive alcohol consumption or tobacco use for both men and women (38), highly educated men are still more likely to engage in risky behaviours and may be less likely to institute healthy behaviour changes compared with highly educated women (39).

As younger physiological age relative to chronological age is associated with reductions in incidence of a range of ageing-related health outcomes, the present analysis highlights how improving gender equity in higher education might reduce gender disparities in healthy ageing by providing women with a midlife buffer to physiological decline. While the gender gap in higher education has been reduced substantially or even reversed in many high-income countries (40), low- and middle-income countries still see considerable gender disparities in education level arising partly from the continued societal expectations of women as homemakers (41). Policies to promote gender equity in education—particularly in settings that still see high levels of gender inequality—might contribute to reducing gender disparities in a wide range of health outcomes in old age.

It is well-established that education has propagating effects on health during the life course (13). The present analysis highlights higher education as particularly important for improving trajectories of physiological ageing in women, possibly reducing the female deficit in health in old age. As the gender gap in higher education is progressively closed, whether women will continue to ‘live longer in worse health’ remains to be seen.

## Data Availability

ELSA data are available to researchers after registration with the UK data service at https://beta.ukdataservice.ac.uk/datacatalogue/series/series?id=200011.

## Author contributions

Conceptualisation: MB, AS.

Methodology: MB, AS.

Validation: AS.

Formal analysis: MB.

Data curation: MB, AS.

Writing –original draft preparation: MB.

Writing –review and editing: MB, AS.

Visualisation: MB.

Supervision: AS. Funding acquisition: AS.

MB and AS had access to all the included data. MB and AS accessed and verified the data. MB had the final responsibility to submit for publication.

## Ethical approval

ELSA Wave 2 received ethical approval from the London Multi-Centre Research Ethics Committee on 12th August 2004 (MREC/04/2/006). ELSA Wave 3 received ethical approval from the London Multi-Centre Research Ethics Committee on 27th October 2005 (05/MRE02/63). ELSA Wave 4 received ethical approval from the National Hospital for Neurology and Neurosurgery & Institute of Neurology Joint Research Ethics Committee on 12th October 2007 (07/H0716/48). ELSA Wave 5 received ethical approval from the Berkshire Research Ethics Committee on 21st December 2009 (09/H0505/124). ELSA Wave 6 received ethical approval from the NRES Committee South Central - Berkshire on 28th November 2012 (11/SC/0374). ELSA Wave 7 received ethical approval from the NRES Committee South Central - Berkshire on 28th November 2013 (13/SC/0532). ELSA Wave 8 received ethical approval from the South Central – Berkshire Research Ethics Committee on 23rd September 2015 (15/SC/0526). ELSA Wave 9 received ethical approval from the South Central – Berkshire Research Ethics Committee on 10th May 2018 (17/SC/0588). ELSA Wave 10 received ethical approval from the South Central – Berkshire Research Ethics Committee on 22nd March 2021 (21/SC/0030). No further ethical approval is required for the present study.

